# Targeting the Histone Methyltransferase SETD7 Rescues Diabetes-induced Impairment of Angiogenic Response by Transcriptional Repression of Semaphorin 3G

**DOI:** 10.1101/2023.12.05.23299540

**Authors:** Shafeeq A. Mohammed, Era Gorica, Mattia Albiero, Gergely Karsai, Alessandro Mengozzi, Carlo Maria Caravaggi, Samuele Ambrosini, Stefano Masi, Maria Cristina Vinci, Gaia Spinetti, Sanjay Rajagopalan, Assam El-Osta, Jaroslav Pelisek, Frank Ruschitzka, Gian Paolo Fadini, Sarah Costantino, Francesco Paneni

**Author notes:** The authors contributed equally to this work (Sarah Costantino, Francesco Paneni). **Address for Correspondence:** Prof. Dr. med. Francesco Paneni, FESC Director, Center for Translational and Experimental Cardiology (CTEC) Head, Cardiometabolic Division University Heart Center Department of Cardiology Rämistrasse 100 CH-8091 Zürich www.herzzentrum.usz.ch.

## Abstract

**Background:** Peripheral artery disease (PAD) is highly prevalent in patients with diabetes (DM) and associates with a poor prognosis. Revascularization strategies failed to improve outcome, suggesting that new strategies to promote blood vessel growth are needed. Histone modifications have emerged as key modulators of gene expression, however their role in angiogenic response in DM remains poorly understood. Here we investigate the role of chromatin remodelling in DM-related impairment of angiogenic response.

**Methodology:** Primary human aortic endothelial cells (HAECs) were exposed to normal glucose (NG, 5 mM) or high glucose (HG, 25 mM) for 48 hours. Gene expression profiling was performed by RNA sequencing (RNA-seq). Cell migration and tube formation were employed to study angiogenic properties in HAECs. Levels of the histone methyltransferase SETD7 and its chromatin signature at histone 3 on lysine 4 (H3K4me1) were investigated by Western blot and chromatin immunoprecipitation (ChIP). Pharmacological blockade of SETD7 was achieved by using the selective inhibitor *(R)*-PFI-2 while the inactive enantiomer (*S*)-PFI-2 was used as a control. Mice with streptozotocin-induced DM were orally treated with (*R*)-PFI-2 or vehicle and underwent hindlimb ischemia by femoral artery ligation. Our experimental findings were translated in endothelial cells and gastrocnemius muscle samples obtained from DM patients with PAD.

**Results:** RNA-seq in HG-treated HAECs unveiled the histone methyltransferase SETD7 as the top-ranking transcript. SETD7 upregulation was associated with increased H3K4me1 levels as well as with impaired HAECs migration and tube formation. Both SETD7 silencing and inhibition by *(R)*PFI-2 rescued hyperglycemia-induced impairment of HAECs migration and tube formation, while SETD7 overexpression blunted the angiogenic response. RNA-seq and ChIP assays showed that SETD7-induced H3K4me1 enables the transcription of the angiogenesis inhibitor semaphorin-3G (SEMA3G) by increasing chromatin accessibility to PPARγ. Moreover, SEMA3G overexpression mimicked the impairment of angiogenic response observed during hyperglycemia. In DM mice with hindlimb ischemia, (*R*)-PFI-2 improved limb perfusion by suppressing SEMA3G. Finally, RNAseq and immunofluorescence in vascular specimens from two cohorts of DM patients with PAD confirmed the upregulation of SETD7/SEMA3G signalling. Of note, (*R*)-PFI-2 restored angiogenic properties in HAECs collected from DM patients.

**Conclusion:** SETD7 is a druggable epigenetic target to promote neovascularization in DM.

## Introduction

Diabetes mellitus (DM) is a major cause of disability due to the occurrence of micro- and macrovascular complications.^1^ Among the constellation of cardiovascular comorbidities, DM shows the highest association with peripheral artery disease (PAD), a condition characterized by atherosclerotic lesions, reduced blood supply and chronic ischemia of the lower extremities.^2^ DM is highly prevalent in patients with PAD and has been co-diagnosed in nearly 40% of all PAD patients.^3^ Despite advances in revascularization strategies, the rate of limb amputation due to chronic limb ischemia remains high and associates with cardiovascular morbidity in DM patients.^3–5^ In this perspective, strategies that promote vascularization can be considered as a novel therapeutic option in DM patients with PAD.^6–8^ Epigenetic changes, defined as plastic chemical modifications of DNA/histone complexes – have shown to modulate gene activity by modifying chromatin accessibility to transcription factors.^9, 10^ To date, only few studies have investigated the contribution of epigenetic modifications in regulating angiogenic response and post-ischemic vascularization.^11, 12^ Inhibition of histone deacetylase 9 (HDAC9) was shown to impair endothelial cells (ECs) tube formation, sprouting and retinal vessel outgrowth, whereas HDAC9 overexpression rescued the impairment of angiogenesis.^13^ In another study, pharmacological inhibition and genetic ablation of transcriptional co-activator p300-CBP associated factor (PCAF) impaired blood flow recovery and arteriogenesis following hindlimb ischemia via suppression of inflammatory signalling and leukocyte recruitment.^14^ Although these studies highlighted a pivotal role of chromatin remodelling in the regulation of EC angiogenic properties, our understanding of epigenetic regulation of angiogenic response in diabetes remains poor and chromatin-editing approaches with specific epi-drugs are yet to be approved in this setting. In the present study we show that, among chromatin modifying enzymes, the histone methyltransferase SETD7 is strongly upregulated by hyperglycemia and impairs angiogenic response by transcriptional regulation of the anti-angiogenic gene semaphorin3G (SEMA3G). Pharmacological targeting of SETD7 by *(R)*PFI-2 was able to rescue DM-induced impairment of angiogenic properties both *in vitro* and *in vivo*. Activation of the SETD7/SEMA-3G axis was validated by RNA-seq and immunofluorescence in two different cohorts of patients with DM and PAD. Notably, SETD7 inhibition by *(R)*PFI-2 rescued migration and tube formation in ECs collected from DM patients thus suggesting the potential clinical relevance of our study. Taken together, these findings suggest that selective pharmacological blockade of SETD7 could help restoring post-ischemic neovascularization thus preventing limb ischemia in DM.

## Methods

An extended version of the methods used in this study is provided in Supplemental Material.

### Experiments in human aortic endothelial cells

Human aortic endothelial cells (HAECs, passages 5 to 7) were cultured in EBM-2 (growth factor-free medium) with 2% FBS and exposed for 48 hours either to normal glucose (5 mmol/L) or high glucose concentrations (25 mmol/L), in the presence or in the absence of the SETD7 inhibitor R-PFI-2 (20 μM) or its inactive enantiomer S-PFI-2 (20 μM). Mannitol at the final concentration of 20 mmol/L was used as an osmotic control. After 48 h cells were harvested and used for angiogenesis assays or molecular analyses.

### RNA-sequencing

RNA-sequencing (RNA-seq) was performed in NG and HG-treated HAECs, in the presence or in the absence of SETD7-siRNA or scrambled siRNA (n=5/group). Samples were sequenced as a75 bp paired end with a NextSeq 500 using the Truseq LT kit (please see Supplemental Material for details about RNA-seq and bioinformatic analysis).

### Chromatin immunoprecipitation (ChIP) assays

Chromatin immunoprecipitation was performed in HAECs and mouse gastrocnemius muscle specimens by using the Magna ChIP Assay Kit (Millipore, Billerica, USA), according to the manufacturer’s instructions. ChIP quantifications of gene promoters were performed by real time PCR (gene primers are shown in **Table S1).** Quantifications were performed using the comparative cycle threshold method and are reported as the n fold difference in antibody-bound chromatin against the input DNA, as previously reported.^15, 16^

### Mouse model of diabetic hindlimb ischemia

Diabetes was induced in 8-week-old male mice (C57BL/6) by a single high dose of streptozotocin (STZ, 175 mg/Kg, via intraperitoneal injection) as previously reported.^17^ For the hindlimb ischemia experiments, animals were sedated with 5% inhaled Isoflurane (Iso-Vet, Piramal Healthcare, UK) and kept at 2-3% for maintenance. Analgesia was achieved with 5 mg/kg tramadol. The femoral artery and the vein were surgically dissected from the femoral nerve, then cauterized by applying an electric current with a bipolar tweezer end excised from the proximal end to popliteal bifurcation. Hindlimb perfusion was measured with Perimed PeriscanPim II Laser Doppler System (PerimedAB, Sweden) at 1, 7 and 14 days after ischemia. Animal experiments were approved by the Veneto Institute of Molecular Medicine Animal Care and Use Committee and by the Italian Health Ministry.

### In vivo treatment with *(R)-*PFI-2

*(R)-*PFI-2 hydrochloride (Cat. HY-18627A, MedChemExpress) at the final dose of 95 mg/Kg or vehicle (DMSO:Corn oil at 1:1 ratio) were given orally in mice for 5 days before the induction of hindlimb ischemia and continued for the next 14 days. Body weight and blood glucose levels were monitored before and after treatment. At the end of the study, mice were harvested, and liver specimens were tested for possible toxicity.

### RNA-seq in DM patients with PAD and healthy control donors

Human vascular tissue samples were collected from DM patients with PAD who underwent open surgical interventions at the Department of Vascular Surgery at the University Hospital Zurich. Control aortas were obtained from healthy donors and provided by the Department of Visceral Surgery at the University Hospital Zurich. The local ethics committee (Cantonal Ethics Committee Zurich, Switzerland; BASEC-Nr. 2020-00378) approved the tissue sample collection and processing for molecular analyses. The detailed methods for library preparation and RNA-seq are reported in Supplemental Material.

### Collection of skeletal muscle specimens from DM patients with PAD

Skeletal muscle specimens were obtained from a second cohort of DM patients with PAD to validate our findings. The collection of human samples was approved by the MultiMedica Research Ethics Committee and was conducted according to the principles outlined in the Declaration of Helsinki. Specimens were collected from: i) different anatomical locations of the lower extremities from non-diabetic control participants, referred for investigations/therapeutic interventions related to leg varicosity; or ii) foot muscle from T2D participants at the occasion of minor amputation for chronic limb ischemia (diagnosed according to the Trans-Atlantic Inter-Society Consensus Document on Management of Peripheral Arterial Disease [TASC].

### Experiments in primary endothelial cells collected from patients

Primary human aortic endothelial cells (passages 5–7, Clonetics®) were obtained from DM patients (M:F=2:2; age, 61±2 years; n=4), free from overt cardiovascular disease and other relevant comorbidities. Cells were grown in fibronectin-coated 75 cm^2^ flasks in optimized endothelial growth medium-2 supplemented with 2% fetal bovine serum, as previously described.^18^ Treatment with *(R)*-PFI-2 was performed as described above and in Supplemental Material.

## Results

### Hyperglycemia leads to an impairment of angiogenic response and SETD7 upregulation in human aortic endothelial cells (HAECs)

To investigate whether HG may impact on angiogenesis *in vitro*, we exposed primary human aortic endothelial cells (HAECs) to normal glucose (NG, 5 mM/L) and high glucose concentrations (HG, 25 mM/L) for 24 and 48 hours. Compared with NG, HG led to an impairment of endothelial migration, tube formation and sprouting **(Fig. 1A-C)**. To unveil transcriptional signatures potentially involved in hyperglycemia-induced impairment of angiogenesis, we employed an unbiased approach by performing RNA-seq in HAECs treated with NG and HG. Among dysregulated genes, HAECs treated with HG displayed a significant upregulation of the histone methyltransferase SETD7 **(Fig. 1D-E; Fig. S1A).** Consistently with this finding, HG led to a time-dependent increase in SETD7 gene expression and protein levels **(Fig. 1F, Fig. S1B-C),** whereas no changes in SETD7 expression were observed with the osmotic control mannitol (**Fig. S1D**). Since SETD7 is specifically involved in mono-methylation of lysine 4 at histone 3 (H3K4me1)^18–20^, we next investigated this chromatin signature and found an increase in H3K4me1 levels in HG-treated HAECs as compared to the NG group **(Fig. 1G).** In line with enhanced histone methylation, we observed an increased nuclear translocation of SETD7 upon HG treatment, as shown by immunoblotting and immunofluorescence **(Fig 1H-I, Fig. S2A-B)**.

**Figure 1.**
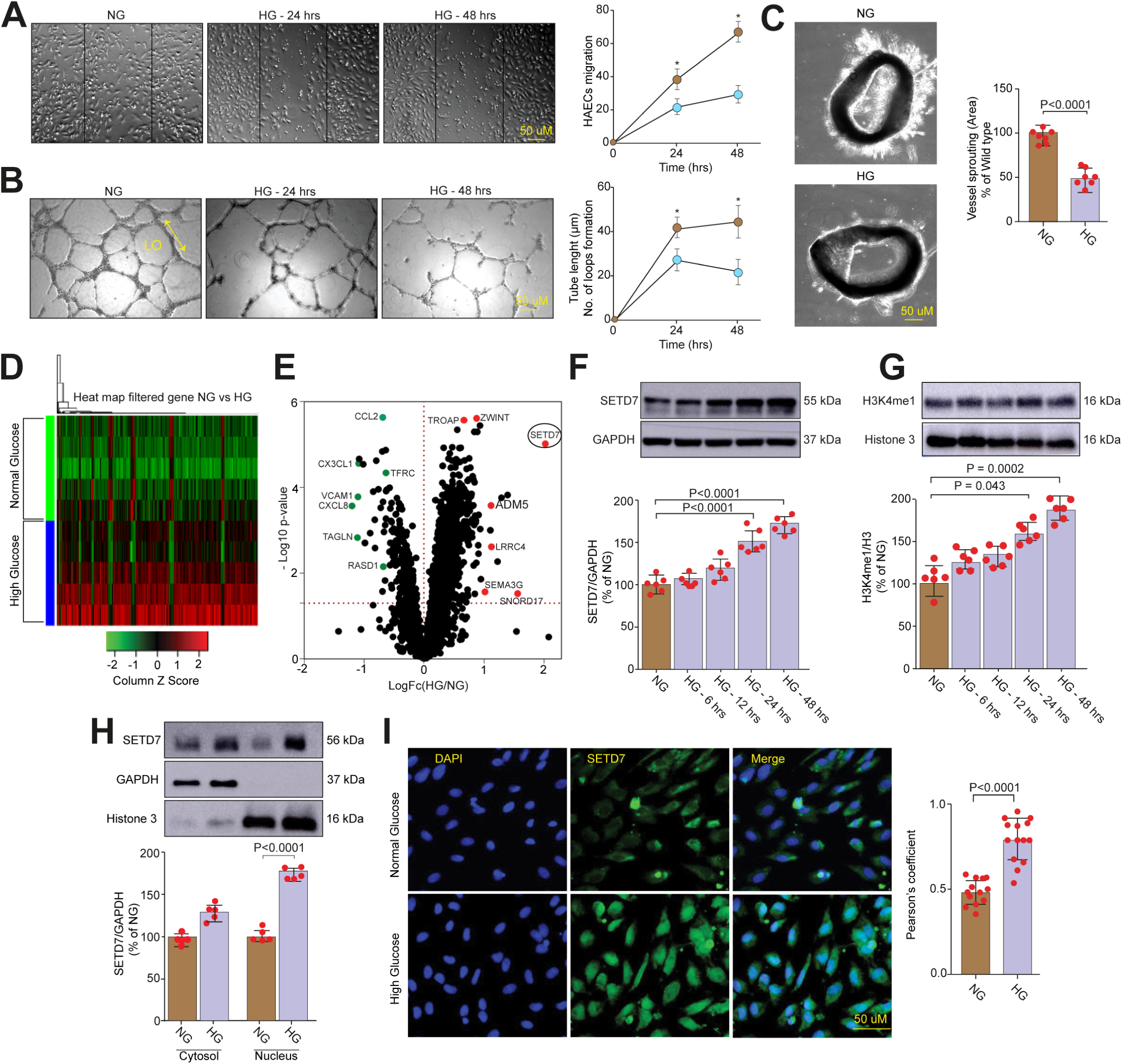
Defective angiogenesis and upregulation of the histone methyltransferase SETD7 by hyperglycemia. HAECs were cultured in growth factor-free medium and exposed to normal glucose concentrations (NG, 5 mM/L) or high glucose concentration (HG, 25 mM/L) for 24 and 48 hours. **A**) Scratch assay and relative quantification showing migration of HAECs exposed to NG and HG; **B**) Representative images of Matrigel-based tube formation assay in the 2 experimental groups. For quantification, the length of tubule (yellow arrow) and loop (LO) formation were considered; **C**) Endothelial sprouting and relative quantification in aortic rings from non-diabetic and diabetic mice; **D-E**) Heatmap and volcano plot displaying differential gene expression in HAECs exposed to NG vs HG; (fold change 2 to -2; significantly downregulated genes are shown in green while upregulated genes are shown in red); **F**) Representative Western blot and relative quantification showing SETD7 protein levels in HAECs treated with NG and HG at different time points (6, 12, 24 and 48 hours). GAPDH and Vinculin were used as loading controls; **G**) Representative Western blot and relative quantification showing H3K4me1 levels in HAECs treated with NG and HG at different time points (6, 12, 24 and 48 hours). Histone 3 was used as a loading control; **H**) Representative Western blot and relative quantification showing SETD7 localization in cytosolic and nuclear fractions from HAECs treated with NG and HG; **I**) Immunostaining and Pearson’s coefficient showing the localization of SETD7 in HAECs treated with NG and HG. Nuclei are shown in blue (DAPI), whereas SETD7 is shown in green. Data are expressed as mean ± SD and shown as a percentage of control. Multiple comparisons were performed by one-way ANOVA followed by Bonferroni post hoc test where appropriate. A p value <0.05 was considered significant. HG, high glucose; NG, normal glucose.

### SETD7 inhibition blunts H3K4me1 levels and rescues hyperglycemia-induced defects of angiogenic response

We next investigated whether SETD7 inhibition could restore angiogenic response in HAECs exposed to HG. Of interest, SETD7 depletion by small interfering RNA (siRNA) reduced H3K4me1 levels while restoring EC migration and tube formation as compared to scrambled siRNA (Scr.siRNA) **(Fig. 2A-C).** Given its specificity for H3K4me1, gene silencing of SETD7 did not affect other H3K4 methylation marks (H3K4me3) **(Fig. S3).** Next, we employed a highly selective pharmacological inhibitor of SETD7, (*R*)-PFI-2, and used its inactive enantiomer (*S*)-PFI-2 as a control. HAECs treated with (*R*)-PFI-2 showed neither signs of cell toxicity nor changes in cell morphology (**Fig. S4A-B**). Consistent with the siRNA experiments, (*R*)-PFI-2 treatment led to a reduction of H3K4me1 levels and associated with an improvement of angiogenic properties (**Fig. 2D-F**). As expected, the drug did not affect H3K4me3 levels (**Fig. S4C**).

**Figure 2.**
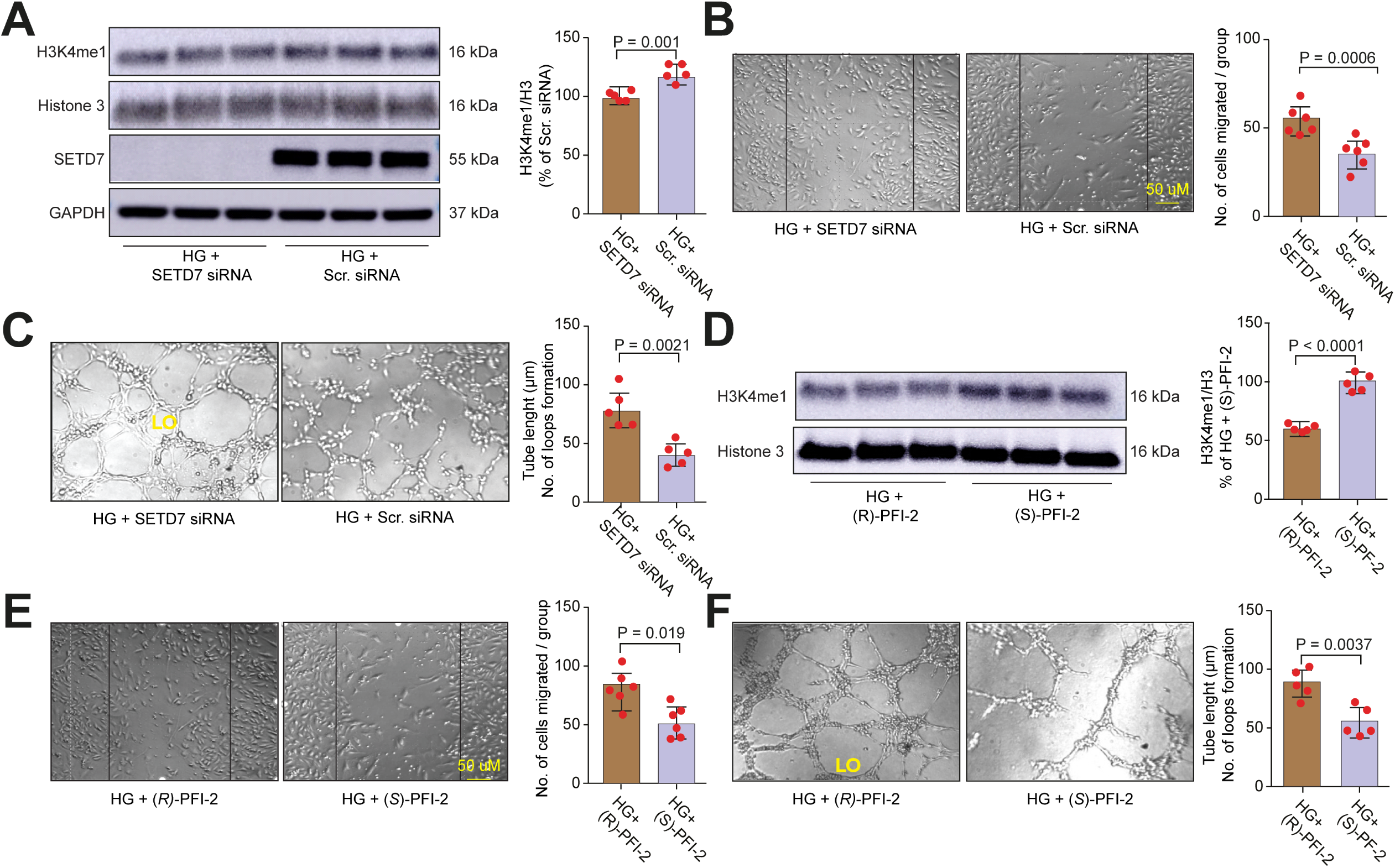
SETD7 inhibition reduces H3K4me1 levels while rescuing HG-induced defects of angiogenic response. **A**) Representative Western blot and relative quantification showing SETD7 and H3K4me1 levels in HAECs treated with HG, in presence of SETD7-siRNA and scramble-siRNA. GAPDH and histone 3 were used as loading controls; **B**) Scratch assay and relative quantification showing migration of HG-treated HAECs in presence of SETD7-siRNA and scramble-siRNA; **C**) Representative images of Matrigel-based tube formation assay in the different experimental groups. For quantification, the length of tubule (indicated by arrow) and loop (LO) formation was considered; **D**) Representative Western blot and relative quantification showing H3K4me1 levels in HAECs treated with HG, in the presence of the SETD7-selective inhibitor *(R)*-PFI-2 and its inactive enantiomer *(S)*-PFI-2. Histone 3 was used as a loading control; **E-F**) Scratch assay and Matrigel-based tube formation assay in HG-treated HAECs treated with *(R)*-PFI-2 or *(S)*-PFI-2. Data are expressed as mean ± SD and shown as a percentage of control. Comparisons were performed by Student’s t test. A p value <0.05 was considered significant.

### SEMA3G is a downstream target of SETD7

We next performed RNA-seq in HG-treated HAECs with and without SETD7 depletion to appraise SETD7-dependent transcriptional changes in ECs (**Fig. 3A**). SETD7 knockdown in HAECs was associated with significant changes in the expression of genes implicated in the regulation of vasculogenesis, angiogenesis and cell migration (as shown by IPA analysis, **Fig. 3B**). Specifically, SETD7-depleted HAECs displayed a significant downregulation of SEMA3G, VCAM1, PTGS1 and DUSP23 (**Fig. 3C, Fig. S5**). In order to unveil SETD7 transcriptional targets, we interrogated the UCSC genome browser (https://genome.ucsc.edu) to investigate the enrichment of SETD7-specific histone marks (H3K4me1) on the promoter of deregulated genes (**Fig. 3D**). SEMA3G promoter showed a strong enrichment in H3K4me1, suggesting a potential involvement of SETD7 in its transcriptional regulation (**Fig. 3D**). SEMA3G was significantly upregulated upon HG exposure as compared to NG (**S6A-B**), while SETD7 gene silencing in HG-treated HAECs led to a significant downregulation of SEMA3G levels as compared to Scr.siRNA, suggesting that SETD7 is required for SEMA3G upregulation (**Fig. 3E, S6A-B**). A similar downregulation of SEMA3G was observed with the selective SETD7 inhibitor (*R*)-PFI-2 as compared to its inactive enantiomer (*S*)-PFI-2 (**Fig. 3F**). On the other hand, SETD7 overexpression in NG-treated HAECs was sufficient to increase SEMA3G protein levels and mimicked hyperglycemia-induced impairment of ECs migration and tube formation (**Fig. 3G-I**).

**Figure 3.**
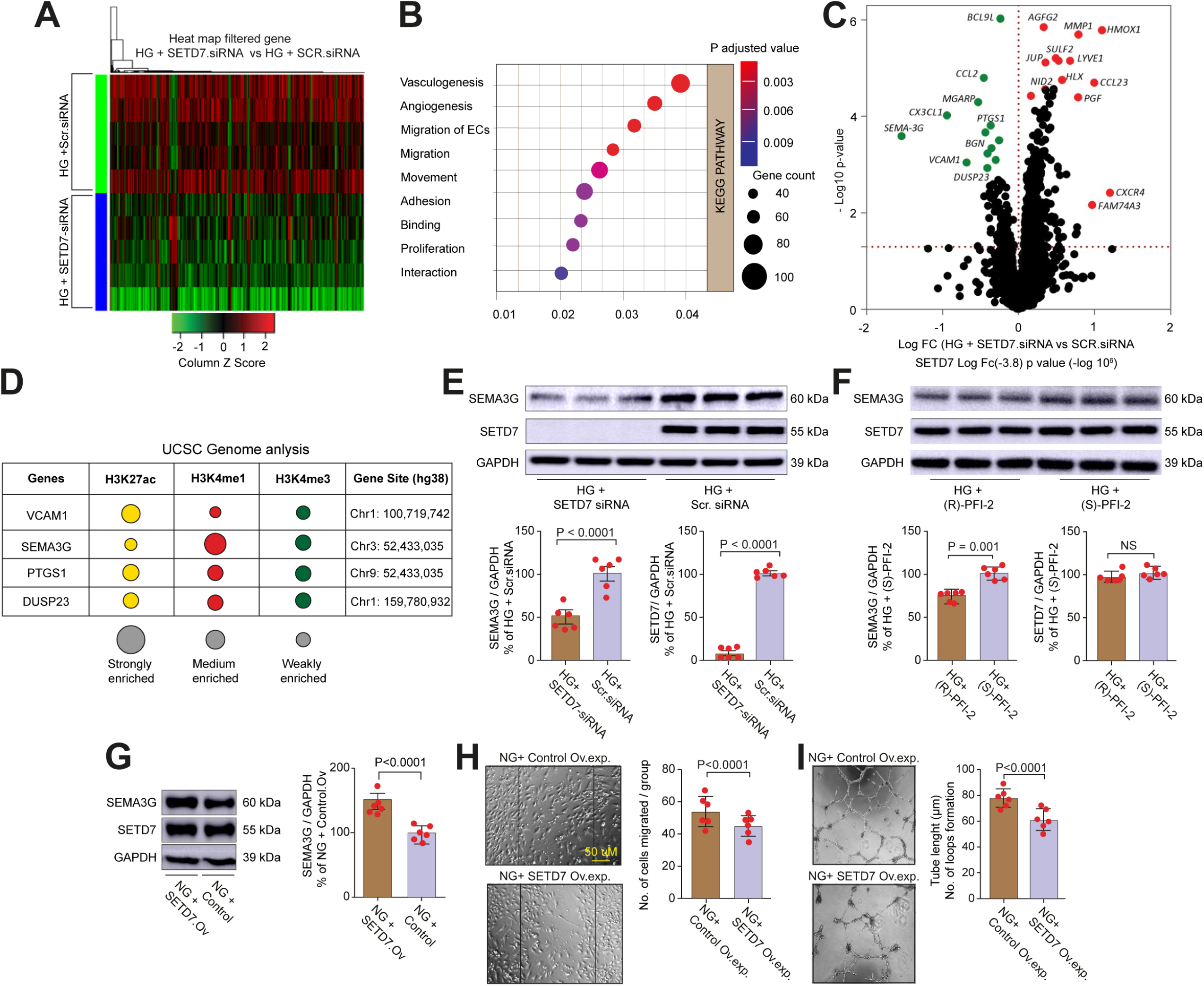
The angiogenesis inhibitor SEMA3G is a downstream target of SETD7. **A**) Heatmap showing differential gene expression in HG-treated HAECs, in the presence of SETD7-siRNA or Scr.siRNA (fold change 2 to -2; downregulated genes are shown in green while upregulated transcripts are shown in red); **B**) IPA (Ingenuity Pathway Analysis) displaying signaling pathways affected by SETD7 depletion; **C**) Volcano plot shows differential gene expression in HG-treated HAECs with and without SETD7 depletion by siRNA (fold change 2 to -2; downregulated genes are shown in green while upregulated transcripts are shown in red); **D**) UCSC genome analysis showing enrichment of different histone marks on gene promoters; **E**) Representative Western blot and relative quantification of SEMA3G in HAECs treated with HG+SETD7 siRNA vs. HG+ Scr.siRNA; **F**) Representative Western blot and relative quantification of SEMA3G in HG-treated HAECs, in presence of the SETD7-selective inhibitor *(R)*-PFI-2 or its inactive enantiomer *(S)*-PFI-2; **G**) Representative Western blot and relative quantification of SEMA3G expression following SETD7 overexpression in HAECs. A scrambled genomic RNA (Scr.Grna) was used as control for SETD7 overexpression; **H**) Scratch assay and relative quantification showing migration of HG-treated HAECs in presence of SETD7 overexpression or Scr.Grna (control); **I**) Representative images of Matrigel-based tube formation assay in the different experimental groups. Multiple comparisons were performed by one-way ANOVA followed by Bonferroni post hoc test where appropriate. A p value <0.05 was considered significant.

### SEMA3G is preferentially expressed in aortic endothelial cells and represses angiogenic capacity

In order to explore the expression pattern of SEMA3G in the vascular endothelium, we analyzed a single-cell RNAseq dataset previously generated in mouse skeletal muscle specimens.^21^ scRNA-seq profiles were obtained from 4,121 cells with a median of 959 genes per cell. Unsupervised clustering followed by t-distributed stochastic neighbor embedding (t-SNE) revealed six clusters of vascular ECs, which all expressed the pan-vascular markers Cdh5 and Pecam1 while did not express described marker sets of muscle stem cells and other muscle cell types.^21^ Of interest, we found that SEMA3G was preferentially enriched in aortic endothelial cells and arteriole endothelial cells (**Fig. 4A**). To characterize the function of SEMA3G in the diabetic endothelium we performed loss-of-function experiments by siRNA. We observed that SEMA3G silencing in HG-treated HAECs restored cell migration and tube formation (**Fig. 4B-C**).

**Figure 4.**
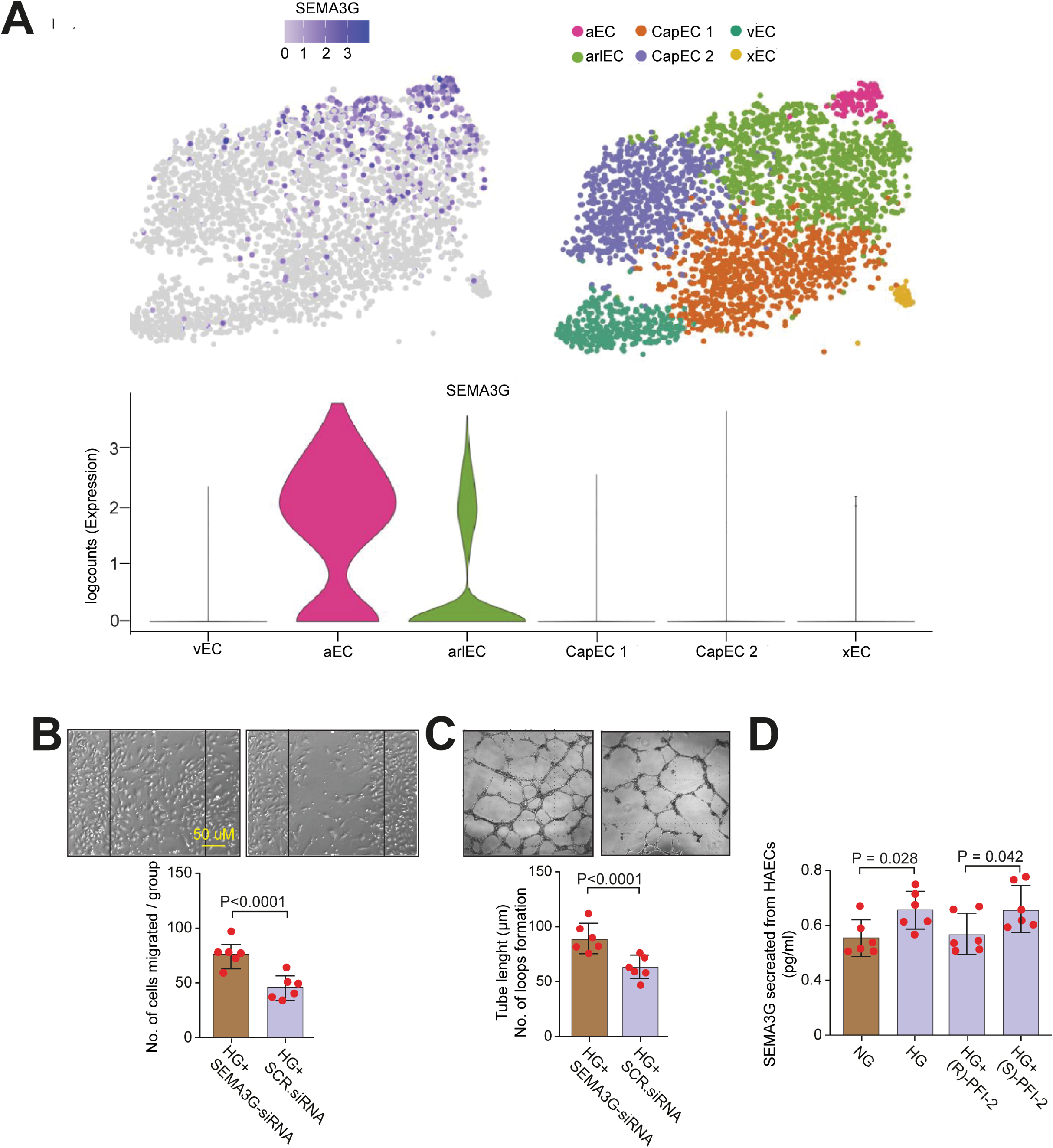
SEMA3G is preferentially expressed in aortic endothelial cells and represses angiogenic capacity. **A)** Bioinformatic analysis of scRNA-seq data obtained in skeletal muscle-derived endothelial cells (https://shiny.debocklab.hest.ethz.ch/Fan-et-al/). Violin plots show normalized SEMA3G expression in each EC cluster; **B)** Scratch assay and relative quantification showing migration of HG-treated HAECs in presence of SEMA3G-siRNA and scr.siRNA; **C)** Representative images of Matrigel-based tube formation assay in the different experimental groups; **D**) ELISA showing SEMA3G levels in conditioned media from HAECs treated with NG and HG, in presence of SETD7-selective inhibitor (R)-PFI-2 and its inactive enantiomer (S)-PFI-2. Multiple comparisons were performed by one-way ANOVA followed by Bonferroni post-hoc test where appropriate. A p value <0.05 was considered significant.

### SETD7 regulates SEMA3G secretion in endothelial cells

Recent work has shown that SEMA3G can be secreted from cells.^22^ Hence, we investigated whether SETD7 inhibition could affect SEMA3G secretion from cultured HAECs exposed to HG. SEMA3G secretion was enhanced in HG-treated HAECs as compared to NG, whereas treatment with the SETD7 inhibitor (*R*)-PFI-2 significantly reduced SEMA3G levels in conditioned media as compared to its inactive enantiomer (*S*)-PFI-2 (**Fig. 4D**).

### SETD7-dependent H3K4me1 favors chromatin accessibility on SEMA3G promoter

To appraise whether SETD7 regulates SEMA3G via H3K4me1 (transcriptional mechanism) rather than by direct protein methylation (posttranslational mechanism), we performed co-immunoprecipitation to test a direct interaction between SETD7 and SEMA3G. No interaction between the 2 proteins was observed upon HG exposure (**Fig. S7**). Bioinformatic analysis of ChIP-seq data from the UCSC genome browser showed a strong enrichment of H3K4me1 on SEMA3G promoter (**Fig. 5A**). To confirm these data, we performed a ChIP assay by leveraging three sets of primers targeting different regions of SEMA3G promoter (**Fig. 5B**). ChIP assay performed in HG-treated HAECs revealed that H3K4me1 is preferentially enriched at the initiator site of SEMA3G promoter (**Fig. S8A**). Interestingly enough, H3K4me1 enrichment on SEMA3G promoter was reduced in the presence of (*R*)-PFI-2 in HG-treated HAECs, as compared to (*S*)-PFI-2 (**Fig. 5C**). Moreover, SETD7 inhibition only affected H3K4me1 but not H3K4me3 enrichment, in line with the notion that SETD7 specifically induces histone mono-methylation (**Fig. S8B**). This set of data shows that SETD7-induced H3K4me1 is required for SEMA3G transcription.

**Figure 5.**
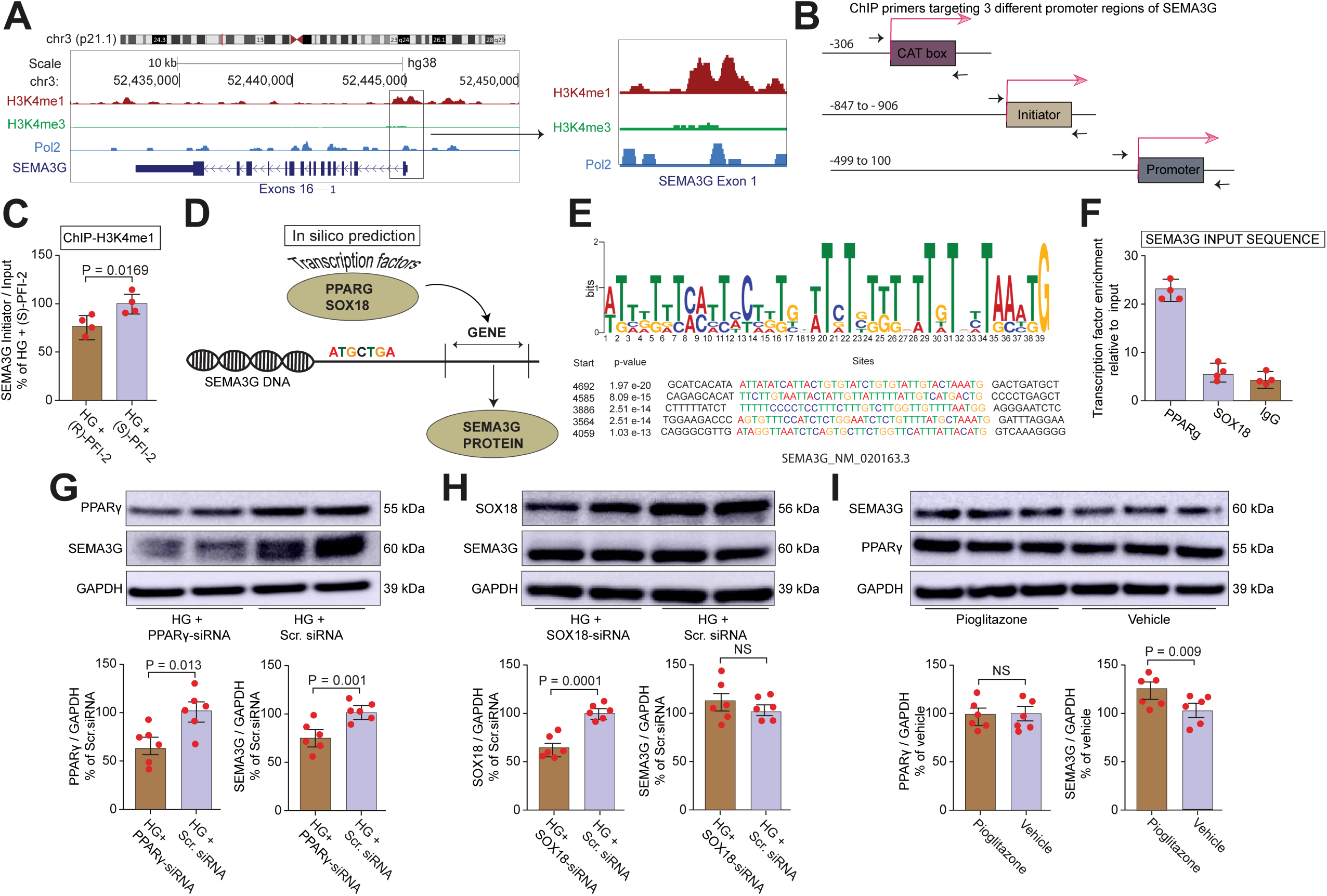
PPARγ regulates SEMA3G transcription. **A**) Analysis of ChIP-seq data (available from the UCSC genome browser) showing enrichment of chromatin marks (H3K4me1, H3K27ac) on SEMA3G promoter; **B**) Schematic showing ChIP primers targeting three different regions of SEMA3G promoter; **C**) ChIP assay showing enrichment of H3K4me1 on SEMA3G promoter in HG-treated HAECs; **D**) *In silico* prediction analysis shows PPARγ and SOX18 as transcription factors potentially involved in SEMA3G transcription; **E**) Motif binding analysis (JASPAR) showing the sequence for transcription binding site; **F**) ChIP assay showing enrichment of PPARγ and SOX-18 on SEMA3G promoter. Chromatin was immunoprecipitated with normal rabbit IgG or antibody against SOX-18 and PPARγ, and precipitated genomic DNA was analyzed by real-time PCR using primers for SEMA3G promoter; n = 4/group; **G**) Representative Western blot and relative quantification of SEMA3G and PPARγ in HG-treated HAECs, in presence of PPARγ siRNA or Scr.siRNA; **H**) Representative Western blot and relative quantification of SEMA3G and SOX18 in HG-treated HAECs with and without SOX18 depletion; **I**) Representative Western blot and relative quantification of SEMA3G and PPARγ expression in HAECs treated with the PPARγ agonist pioglitazone or vehicle (DMSO). Multiple comparisons were performed by one-way ANOVA followed by Bonferroni post-hoc test where appropriate. A *p* value <0.05 was considered significant.

### PPARγ drives SEMA3G transcription

*In silico* analysis showed that both PPARγ and SOX18 could potentially act as transcription factors on the SEMA3G promoter (**Fig. 5D**). By leveraging motif binding analysis (https://jaspar.genereg.net/) we next determined the nucleotide sequence of transcription factor binding site (**Fig. 5E**). ChIP assay revealed a strong binding of the transcription factor PPARγ to SEMA3G promoter, while no binding was observed for SOX18 (**Fig. 5F**). These results were confirmed by experiments with siRNA-mediated depletion of both PPARγ and SOX18 showing a reduction of SEMA3G expression and protein levels only in presence of PPARγ but not SOX18-siRNA (**Fig. 5G-H, Fig. S9A-B**). PPARγ-siRNA did not affect SETD7 protein levels (**Fig. S9C**). To further validate the role of PPARγ in SEMA3G transcription, we treated HAECs with the PPARγ agonist pioglitazone. Notably, HAECs treated with pioglitazone showed increased SEMA3G expression and protein levels as compared to vehicle (**Fig. 5I, Fig. S10A-B**). Pioglitazone did not affect SETD7 expression (**Fig. S10B-C**).

### (*R*)-PFI-2 improves post-ischemic vascularization and limb perfusion in diabetic mice

Prompted by our *in vitro* results, we next determined whether SETD7 inhibition could rescue ischemia-mediated angiogenesis *in vivo*, in a model of diabetic hindlimb ischemia. Diabetes was induced by streptozotocin while control mice received citrate buffer alone, as previously reported.^17^ After 4 weeks, subgroups of diabetic and non-diabetic mice were randomized to oral treatment with (*R*)-PFI-2 (95 mg/kg/day) or vehicle 6 days before the induction of hindlimb ischemia till 14 days following limb ischemia, for a total of 20 days. Alanine transaminase (ALT) assay excluded liver toxicity in mice treated with (*R*)-PFI-2 (**Fig. S11A**). Diabetes led to increased blood glucose levels and body weight loss as compared to non-diabetic mice, and these alterations were not affected by *(R)*-PFI-2 treatment (**Fig. S11B-C**). Following hindlimb ischemia, diabetic mice exhibited a marked impairment of blood perfusion as assessed by laser Doppler Imaging (blood flow of ischemic vs. non-ischemic hindlimb at 14 days (0.48 ±0.02 vs 0.36 ± 0.02, *p* = 0.003) as compared to non-diabetic controls (**Fig.6A-B**). Treatment with (*R*)-PFI-2 restored blood perfusion levels in diabetic mice as compared to vehicle (0.36 ± 0.02 vs 0.51 ± 0.04, *p*=0.025) (**Fig. 6A-B**). The observed improvement of limb perfusion with (*R*)-PFI-2 was associated with restoration of capillary density, as shown by CD31 immunofluorescence in cross-sections from gastrocnemius muscles of diabetic mice (**Fig. 6D**).

**Fig. 6.**
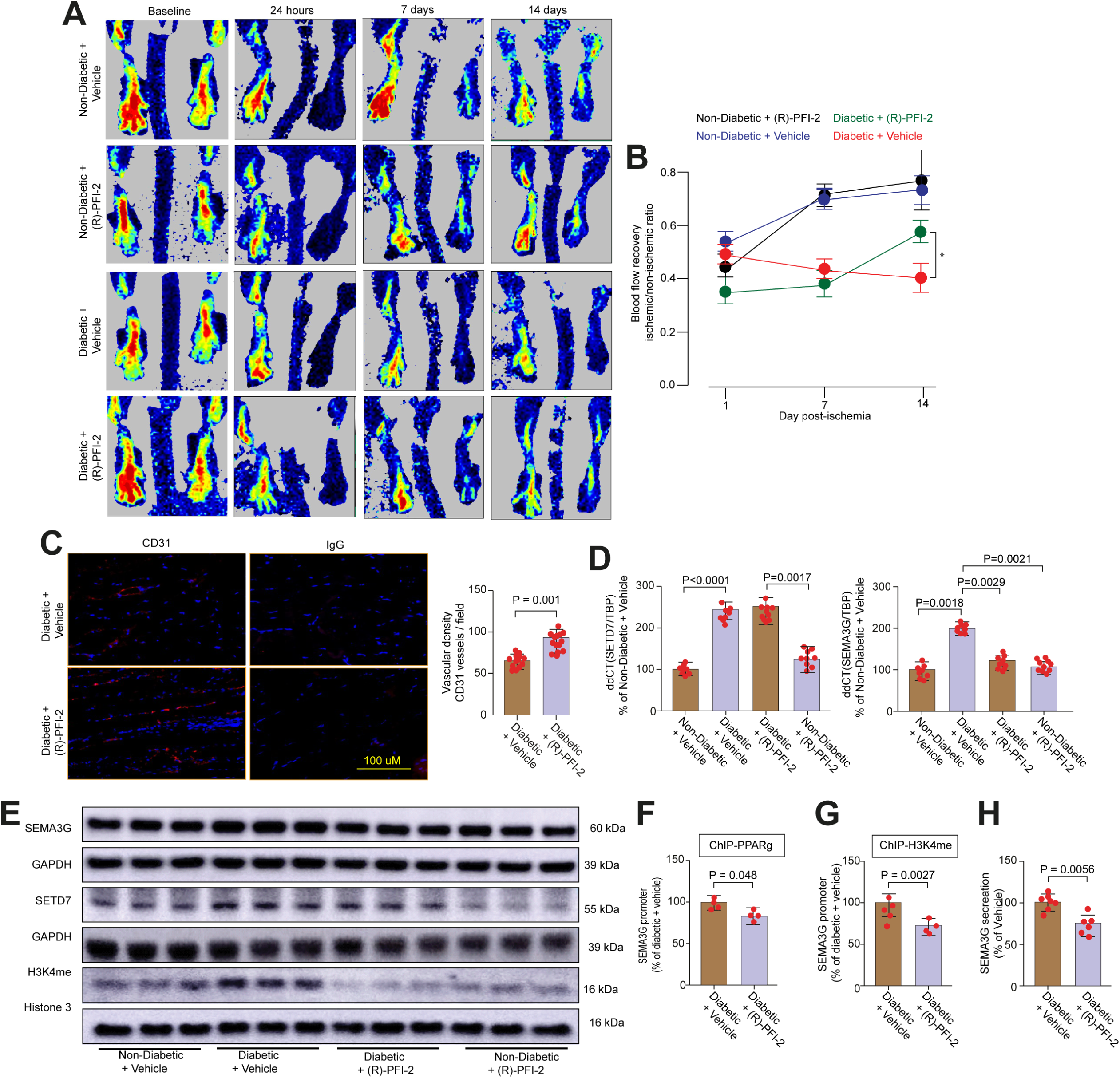
SETD7 inhibition by (*R*)-PFI-2 improves post-ischemic vascularization and limb perfusion in diabetic mice. Unilateral hindlimb ischemia was surgically induced in non-diabetic and diabetic C57BL/6J male mice; **A**) Laser Doppler perfusion imaging was serially performed to determine blood flow recovery at 24 h, 7 and 14 days after hindlimb ischemia; **B**) Time-dependent changes of blood flow recovery (expressed as the ratio of blood perfusion in the ischemic versus nonischemic hind limb) in diabetic and non-diabetic mice treated with (*R*)-PFI-2 or vehicle; *, p<0.05; **C**) Immunostaining (CD31) and relative quantification showing capillary density in gastrocnemius muscle samples. Scale bar = 50 μm; n = 9/group; **D)** Real-time PCR showing gene expression of SETD7 and SEMA3G in gastrocnemius muscle obtained from non-diabetic and diabetic mice treated with vehicle or (R)-PFI-2; **E)** Representative Western blot of STED7, SEMA3G and H3K4me1 in gastrocnemius muscle obtained from non-diabetic mice and diabetic mice treated with vehicle or *(R)*-PFI-2. GAPDH and histone 3 were used as loading controls**; F-G)** ChIP assay showing enrichment of PPARG and H3K4me1 on SEMA3G promoter. Chromatin was immunoprecipitated with normal rabbit IgG or antibody against PPARG and H3K4me1, and precipitated genomic DNA was analyzed by real-time PCR using primers for SEMA3G promoter, n=4/group. **H)** Circulating SEMA3G levels in plasma samples from diabetic mice treated with (*R*)-PFI-2 or vehicle. Data are expressed as mean ± SD and shown as a percentage of control. Multiple comparisons were performed by one-way ANOVA followed by Bonferroni correction and Student’s t-test where appropriate. A p value <0.05 was considered significant.

### In vivo modulation of SETD7/SEMA3G axis by (*R*)-PFI-2

In line with our *in vitro* data, SETD7/SEMA3G axis was dysregulated in ischemic gastrocnemius muscles from diabetic as compared to non-diabetic mice, as shown by real time PCR and Western blot (**Fig. 6E, Fig. S12**). (*R*)-PFI-2 treatment suppressed SETD7/SEMA3G expression while decreasing H3K4me1 levels (**Fig. 6E, Fig. S12**). Mechanistically, ChIP assays showed that (*R*)-PFI-2 prevented the enrichment of both H3K4me1 and the transcription factor PPARγ on SEMA3G promoter (**Fig. 6F-G).** Of interest, we found that in vivo treatment with (*R*)-PFI-2 reduced plasma levels of SEMA3G in diabetic mice (**Fig. 6H**).

### SETD7/SEMA3G signaling in diabetic patients with PAD

To translate our experimental findings to humans, we first performed RNA-seq in vascular specimens collected from DM patients with PAD undergoing elective surgical revascularization and age-matched healthy controls. Of note, RNA-seq confirmed a significant dysregulation of both SETD7 and SEMA3G (**Fig. 7A**). SETD7/SEMA3G axis and H3K4me1 expression were then validated in a second cohort of patients, where gastrocnemius muscle specimens were collected from non-diabetic patients (controls) and age-matched patients with DM and PAD. Immunofluorescence confirmed the upregulation of SETD7, SEMA3G and H3K4me1 in DM patients as compared to controls (**Fig.7B-C**). Pearson’s correlation confirmed the nuclear translocation of SETD7 in muscular specimens from DM patients as compared to controls (**Fig. 7D**). Real-time PCR confirmed SETD7 and SEMA3G upregulation in DM patients with PAD **(Fig. 7E**). Interestingly, ChIP assay confirmed the enrichment of PPARγ and H3K4me1 on SEMA3G promoter in muscle specimens from DM patients with PAD as compared to controls (**Fig. 7F-G**).

**Fig. 7.**
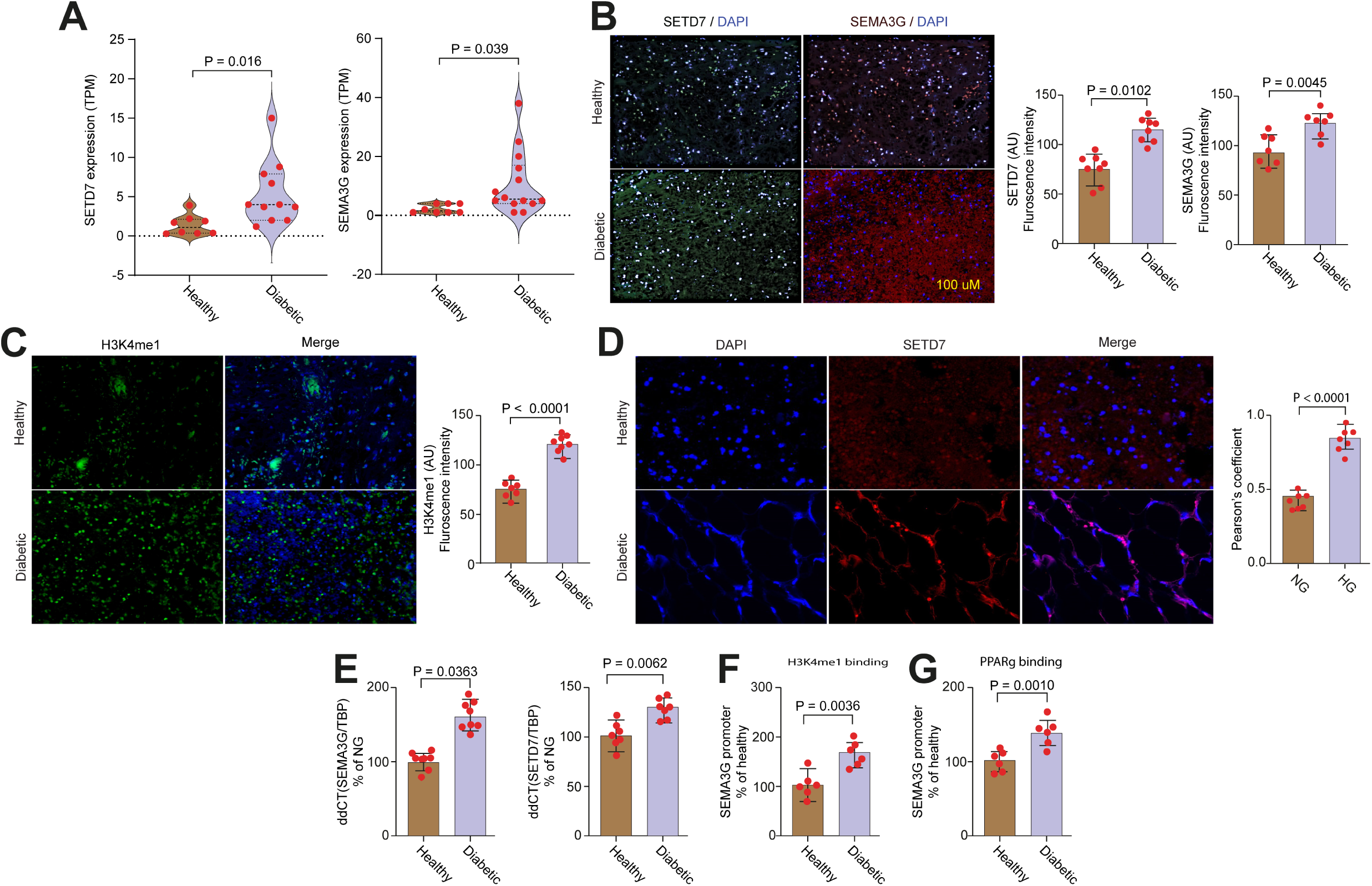
Dysregulation of SETD7/SEMA3G signaling in patients with DM and PAD. **A)** RNA-seq data show the upregulation of SETD7 and SEMA3G gene expression in vascular specimens from DM patients with PAD as compared to healthy controls; **B-C**). Immunofluorescence and relative quantification showing SETD7, SEMA3G and H3K4me1 expression in lower limb muscle specimens from healthy and DM patients with PAD; **D**) Representative immunofluorescence images showing localization of SETD7 in lower limb muscle specimens from DM patients with PAD and non-diabetic controls. SETD7 displays a cytosolic localization in non-diabetic patients while it shows nuclear translocation in DM patients with PAD. Nuclei were stained with DAPI (blue). Quantification of SETD7 nuclear translocation was performed by using the Pearson’s coefficient; **E**) Real-time PCR confirming theupregulation of SEMA3G and SETD7 in muscular specimens from DM patients with PAD versus non-diabetic controls; **F-G**) ChIP assay showing enrichment of H3K4me1 and PPARγ on SEMA3G promoter in muscular specimens from DM patients with PAD versus non-diabetic controls. Chromatin was immunoprecipitated with normal rabbit IgG or antibody against PPARΓ and H3K4me1, and precipitated genomic DNA was analyzed by real-time PCR using primer of SEMA3G promoter. Data are expressed as mean ± SD and shown as a percentage of control. Multiple comparisons were performed by one-way ANOVA followed by Bonferroni correction and Student’s t-test where appropriate. A p value <0.05 was considered significant. *p < 0.05; **p < 0.01; and ***p < 0.001. PAD, peripheral artery disease; T2D, type 2 diabetes.

### Targeting SETD7 restores angiogenic properties in the human diabetic endothelium

We next investigated whether pharmacological modulation of SETD7 in the human diabetic endothelium could rescue angiogenic properties. To this end, we employed human aortic endothelial cells isolated from patients with DM (M:F=2:2; age, 61±2 years; n=4). (*R*)-PFI-2 attenuated SEMA3G expression and H4K3me1 levels in diabetic ECs as compared to its inactive enantiomer (*S*)-PFI-2 (**Fig. 8A-B**). Moreover, SETD7 inhibition significantly improved EC cell migration and tube formation (**Fig. 8C-D**). A schematic summarizing the main study findings is reported in **Fig. 8E**.

**Fig. 8.**
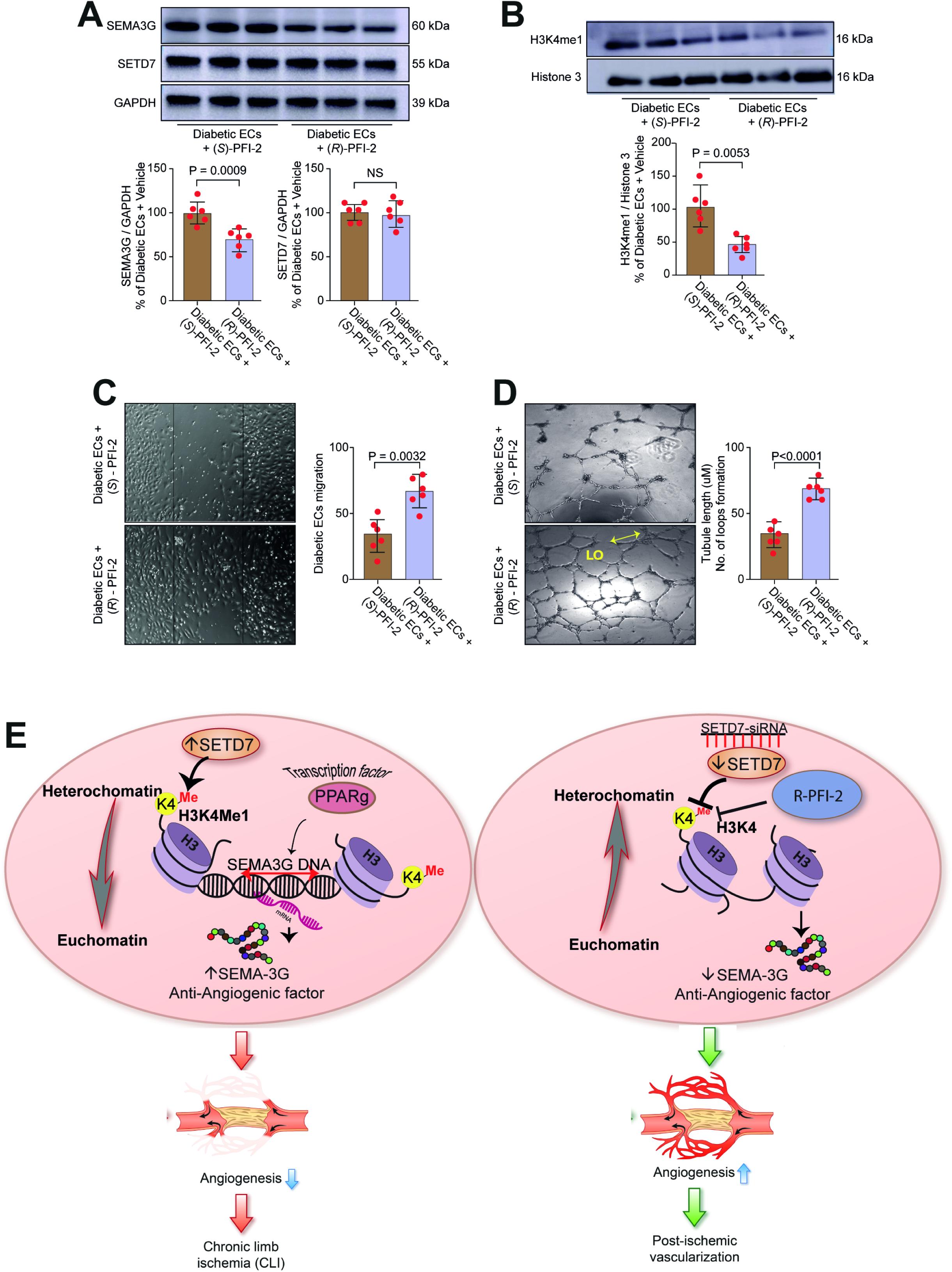
(*R*)-PFI-2 restores angiogenic properties in the human diabetic endothelium. **A-B)** (*R*)-PFI-2 attenuates SEMA3G expression and H4K3me1 levels in human aortic endothelial cells isolated from patients with DM (D-HAECs). The inactive enantiomer *(S)*-PFI-2 was used as a negative control; **C-D**) Cell migration and tube formation in D-HAECs treated with (*R*)-PFI-2 and *(S)*-PFI-2. Comparisons were performed by Student’s t-test where appropriate. A p value <0.05 was considered significant; **E**) Schematic showing the role of SETD7/SEMA3G axis and the effects of its therapeutic modulation to restore vascularization in diabetic PAD.

## Discussion

In the present study we show that, among different genes deregulated by hyperglycemia, the histone methyltransferase SETD7 is a pivotal chromatin remodeler fostering transcriptional programs leading to defective angiogenesis and reduced limb perfusion in diabetic mice. The most important findings of our study are: i) RNA-seq in human aortic endothelial cells exposed to hyperglycemia unveiled SETD7 as the top-ranking transcript; ii) hyperglycemia increases SETD7 expression and SETD7-dependent H3K4me1, thus leading to an open chromatin and active transcription of the anti-angiogenic gene SEMA3G; iii) Both gene silencing and selective pharmacological inhibition of SETD7 by (*R*)-PFI-2 blunt H3K4me1 levels and SEMA3G transcription thus rescuing hyperglycemia-induced impairment of angiogenic properties; iv) in diabetic mice, the SETD7 inhibitor (*R*)-PFI-2 rescues post-ischemic vascularization and limb perfusion by suppressing SEMA3G transcription; (v) SETD7/SEMA3G signalling was dysregulated in two different cohorts of DM patients with PAD; iv) treatment with (*R*)-PFI-2 in ECs collected from DM patients restored angiogenic properties.

Over the last few years, an increasing body of evidence showed that epigenetic changes affect transcriptional programs implicated in blood vessel growth and angiogenic response.^23^ The growing understanding of chromatin structure has led to the design of specific drugs with ability to erase or write epigenetic signatures eventually resetting the cell transcriptome in disease states.^20^ Notably, several of these compounds have been already approved by the FDA for the treatment of cancer, neurological and autoimmune diseases.^24^ Post-translational modifications of histone tails, namely acetylation and methylation, induce chromatin remodelling that may either enable or repress gene transcription.^10^ *In vitro* studies have recently shown that SETD7 is involved in epigenetic regulation of the transcription factor NF-kB.^25, 26^ In addition, SETD7 induces MCP-1 upregulation and endoplasmic reticulum stress in the kidney of diabetic mice.^27^ Although these studies provide important mechanistic insights on the role of SETD7 in hyperglycemia-induced inflammation, SETD7 function in the vascular endothelium remains poorly understood.

In the present study, we show for the first time that SETD7 is a pivotal repressor of blood vessel growth in the setting of diabetic PAD. Specifically, in conditions of hyperglycaemia SETD7 monomethylates H3K4me1 in proximity of SEMA3G promoter. These chromatin signatures enables changes in chromatin accessibility ultimately favouring SEMA3G transcription and subsequent impairment of EC migration and tube formation. This was demonstrated by SETD7 gain- and loss-of-function approaches. SETD7 overexpression mimicked defective angiogenesis while its depletion or pharmacological blockade by (*R*)-PFI-2 rescued hyperglycaemia-induced impairment of angiogenic properties. We showed that SETD7-dependent histone mono-methylation was causally involved in SEMA3G upregulation. First, co-immunoprecipitation experiments ruled out a direct protein methylation of SEMA3G by SETD7. Furthermore, SETD7 is the only methyl-writing enzyme responsible for H3K4me1^19^ and this confers specificity to our findings. Indeed, we did not observe any change of other H4K4 methylation sites (i.e.H3K4me3) following SETD7 manipulation. However, H3K4me1 is amenable to demethylation by the lysine-specific demethylase 1 (LSD1)^26, 28^ and further experiments should elucidate the fine balance and interplay between SETD7 and LSD1 in the regulation of SEMA3G transcription. An active downregulation of LSD1 could also participate to enhanced H3K4me1 levels in our study.

Although histone methylation is emerging as a pivotal determinant of transcriptional and phenotypic changes in different cell types^29, 30^, methylation of non-histone proteins by SETD7 may also play a role in endothelial homeostasis. For example, SETD7-dependent methylation and LSD1-dependent demethylation of hypoxia-inducible factor-1α (HIF-1α) were recently reported to regulate protein stability in the nucleus in a proline hydroxylation- and VHL-independent manner during normoxic and hypoxic conditions.^31^ We cannot exclude that non-histone methylation by SETD7 could participate to the observed changes in angiogenic properties, and that other SETD7 targets could be involved in the setting of DM.. An interesting aspect of our study is that, once transcribed, SEMA3G is secreted by endothelial cells, and this process is regulated by SETD7. Indeed, both SETD7 depletion by siRNA and pharmacological blockade by (*R*)-PFI-2 were able to blunt SEMA3G levels in conditioned media from cultured human endothelial cells. Of note, circulating levels of SEMA3G were increased in diabetic mice with hindlimb ischemia, while in vivo treatment with (*R*)-PFI-2 led to a significant reduction of SEMA3G levels in plasma. Hence, SETD7 inhibition could prevent detrimental actions of SEMA3G in target organs. In this regard, a recent study showed that SEMA3G promotes adipocyte differentiation, adipogenesis and insulin resistance while its inhibition prevented metabolic alterations in mice.^32^ Moreover, SEMA3G plasma levels were increased in obese patients and positively correlated with circulating leptin and adipokines levels.^32^ Hence, SETD7 inhibition could contribute to prevent SEMA3G-dependent metabolic alterations thus broadening the potential applicability of our findings. Furthermore, our results indicate that SEMA3G could represent a potential biomarker of defective angiogenesis, and future studies should explore this possibility.

In our work, we identified PPARγ as the transcription factor responsible for SEMA3G transcription. ChIP experiments confirmed PPARγ binding to SEMA3G promoter while PPARγ depletion prevented SEMA3G transcription in HAECs. These findings were further confirmed by treatment with the PPARγ agonist pioglitazone which was sufficient to induce SEMA3G transcription. These data could also help to understand the results of clinical trials where pioglitazone was associated with increased hazard for surgical or percutaneous lower extremity revascularization in patients with DM.^33^ In line with our findings, a previous study showed that *in vivo* treatment with pioglitazone was associated with decreased ischemic limb perfusion and capillary density in a rat model of hindlimb ischemia.^34^ Hence, pioglitazone-induced SEMA3G could be a plausible mechanistic explanation for the observed impairment of limb perfusion with this drug.

Consistent with our *in vitro* observations, *in vivo* inhibition of SETD7 by (*R*)-PFI-2 was able to restore limb perfusion and capillary density in diabetic mice with hindlimb ischemia. At the molecular level, we showed that oral administration of (*R*)-PFI-2 was associated with blunted H3K4me1 levels as well as reduced SEMA3G transcription and secretion in plasma. The SETD7 pharmacological inhibitor *(R)*-PFI-2 was recently reported as a first-in-class, potent (Kiapp = 0.33 nM), selective, and cell-active inhibitor of the SETD7 methyltransferase activity.^35, 36^ Our results in diabetic mice pave the way for translational studies testing the effects of SETD7 inhibitors in preclinical models of PAD and limb ischemia. To appraise whether our experimental findings hold true in the human setting, we further investigated SETD7/SEMA3G signalling in two different cohorts of diabetic patients. RNA-seq and immunofluorescence confirmed the upregulation of SETD7/SEMA3G axis in diabetic patients with PAD as compared to healthy controls. Furthermore, we showed a significant enrichment of H3K4me1 on SEMA3G promoter in patient samples, suggesting that (R)-PFI-2 might represent a promising therapeutic intervention in this clinical setting. Of clinical relevance, *(R)*-PFI-2 restored angiogenic properties in ECs obtained from DM patients, suggesting the possibility of a therapeutic modulation of SETD7 in DM patients. The lack of toxicity and adverse effects associated with *(R)*-PFI-2 treatment in mice supports the plausibility of testing *(R)*-PFI-2 in a pre-clinical and clinical setting.

Our study has some limitations. Although hyperglycemia was able to induce a significant upregulation of SETD7, the mechanistic link between high glucose and increased SETD7 activity remains to be elucidated. The N-terminal region of SETD7 contains membrane occupation and recognition nexus (MORN) motifs, which are known to be involved in stress response and regulation of SETD7 localization.^37^ Further experiments should elucidate the role of MORN motifs as a potential mechanism linking cellular stress to SETD7 localization and activity in our setting. Furthermore, the role of other cells potentially involved in the in vivo improvement of limb perfusion and capillary density was not addressed in this study. Although we show an important function of SETD7 in endothelial cells, other relevant cell types, namely pericytes and immune cells, could contribute to the observed phenotype.

In conclusion, our translational study unveils a druggable target to boost post-ischemic vascularization in the setting of diabetes. Our results pave the way for preclinical studies in larger animal models to better define the potential of SETD7-targeting approaches in DM patients with PAD and limb ischemia.

## Data Availability

The data that support the findings of this study are available from the corresponding author upon reasonable request.

https://genome.ucsc.edu

https://shiny.debocklab.hest.ethz.ch/Fan-et-al/

## Sources of Funding

This work was supported by the Swiss National Science Foundation (n. 310030_197557), the Swiss Heart Foundation (n. FF19045), the Olga Mayenfisch Foundation, the Swiss Life Foundation, the Kurt und Senta-Hermann Stiftung, the EMDO Stiftung, the Schweizerische Diabetes-Stiftung, the Novo Nordisk Foundation and the Novartis Foundation for Biomedical Research (to F.P.); the Holcim Foundation and the Swiss Heart Foundation (to SC); the Italian Ministry of Health (Ricerca Corrente to the IRCCS MultiMedica).

## Disclosures

There are no competing interests related to the present work. F.P. is a consultant to Vectura Fertin Pharma and has received personal fees for lectures from Novo Nordisk.

## Abbreviations

PAD: Peripheral artery disease
CLI: Critical limb ischemia
DM: Diabetes mellitus
CAD: Coronary artery disease
EC: Endothelial cell
HAECs: Human aortic endothelial cells
NG: Normal Glucose
HG: High Glucose
ChIP: Chromatin immunoprecipitation
VEGF: Vascular endothelial growth factor
H3k27ac: Histone 3 lysine 27 acetylation
MMT: 3-(4,5-dimethylthiazol-2-yl)-2,5-diphenyltetrazolium bromide
PCR: Polymerase chain reaction
SDS: sodium dodecyl sulfate
siRNA: small interfering RNA
GAPDH: Glyceraldehyde 3-phopsphate dehydrogenase
STZ: streptozotocin
T2D: type 2 diabetes
TBP: TATA-Box binding protein
ChIP: Chromatin immunoprecipitation
ANOVA: analysis of variance
FDR: False discovery rate

## Notes

### Competing Interest Statement

The authors have declared no competing interest.

### Clinical Trial

N/A

### Author Declarations

The local ethics committee (Cantonal Ethics Committee Zurich, Switzerland; BASEC-Nr. 2020-00378) approved the tissue sample collection and processing for molecular analyses. The collection of human samples was approved by the MultiMedica Research Ethics Committee and was conducted according to the principles outlined in the Declaration of Helsinki.

